# Analysis of 470,000 exome-sequenced UK Biobank participants identifies genes containing rare variants which confer dementia risk

**DOI:** 10.1101/2024.11.25.24317885

**Authors:** Lily Gibbons, David Curtis

**Author notes:** Correspondence: David Curtis, UCL Genetics Institute, University College London, Darwin Building, Gower Street, London WC1E 6BT, United Kingdom.

## Abstract

**Background:** Previous studies have reported that rare coding variants in a handful of genes have major effects on risk of Alzheimer’s disease (AD). A recent exome wide association study (ExWAS) of dementia in a subset of the UK Biobank cohort implicated a number of genes, including five which were novel. Here we report a similar analysis, carried out on the full cohort of 470,000 exome-sequenced participants.

**Methods:** A score was assigned to each participant depending on individual and/or parental diagnosis of dementia. Regression analysis including *APOE* ε3 and ε4 doses as covariates was applied to gene-wise tests for association with loss of function (LOF) and nonsynonymous variants. 45 tests using different pathogenicity predictors were applied to the first cohort of 200,000 participants. Subsequently the 100 genes showing strongest evidence for association were analysed in the second cohort of 270,000 participants, using only the best-performing predictor for each gene.

**Results:** Three genes achieved statistical significance, *TREM2, SORL1* and *ABCA7*. The five genes reported as novel in the ExWAS did not produce any appreciable evidence for association in this study. The effects and frequencies of variants in different functional categories were characterised for these genes.

**Conclusions:** Rare coding variants in a small number of genes have important effects on dementia risk. Further study of individual variant effects might elucidate mechanisms of pathogenesis. Incorporating rare variant effects for individual risk assessment might become important if preventative treatments for dementia become available.

## Introduction

Dementia can result from a number of different pathological processes, with the bulk of cases being due to Alzheimer’s disease (AD) or vascular dementia (VD) (Rizzi et al., 2014). However distinguishing the underlying cause can require intensive investigations so that in broadly focused epidemiological studies one may be obliged to consider the phenotype of interest to be undifferentiated dementia. A number of risk factors can contribute to these different underlying causes and these disease-specific risk factors may then be seen to be associated with the overall risk of dementia (Mundada et al., 2024). Although genetic risk factors modifying cardiovascular risk are associated with VD, the clearest associations with undifferentiated dementia are for genes influencing susceptibility to AD (Caplan et al., 2023; Kjeldsen & Frikke-Schmidt, 2024; Latimer et al., 2021). Rare variants in *APP, PSEN1* and *PSEN2* are causally involved in early onset AD while rare variants in *PSEN1* have also been shown to be associated with the common, late onset form of AD (Curtis et al., 2019; Latimer et al., 2021). Other studies have identified rare variants in *TREM2, SORL1, ABCA7, ATP8B4* and *ABCA1* as also being implicated in AD susceptibility (Holstege et al., 2022). Most recently, an exome-wide association study (ExWAS) in the UK Biobank cohort using a phenotype described as clinically diagnosed/proxy AD and related dementia (ADRD) applied gene-based analyses to identify associations for eleven genes significant after FDR correction: S*ORL1, GRN, PSEN1, ABCA7, GBA, ADAM10, FRMD8, DDX1, DNMT3L, TGM2* and *MORC1* (Zhang et al., 2024).

Here we describe a study based on the same exome-sequenced UK Biobank cohort as used in this ExWAS which is in some ways similar but which differs from it in a number of important respects, listed as follows.

The ExWAS was restricted to only White British participants whereas our study uses all available data. We have previously shown that the methods we utilise can deal satisfactorily with ancestrally heterogeneous cohorts (Curtis, 2020).

The ExWAS used a dichotomous phenotype, taking as a case anybody with a personal diagnosis of dementia or with a parent or sibling with dementia. Instead, we have constructed a dementia risk score, which is higher for people with a personal diagnosis of dementia or with two affected parents than for people with just a single affected parent.

The ExWAS carried out multiple analyses for each gene, using different pathogenicity and frequency based masks to group together different sets of variants to be included in collapsing analyses. By contrast, we weighted all variants by frequency and incorporate nonsynonymous and loss of function (LOF) separately in single gene-based analyses, an approach which we have shown can have more power than simple collapsing analyses (Curtis, 2022).

The ExWAS used only a single predictor, REVEL, to assess the pathogenicity of nonsynonymous variants (Ioannidis et al., 2016). However we have shown that different prediction methods vary in their performance across different genes and phenotypes, meaning that no single method is always optimal and in the trials we performed REVEL did not on average outperform other methods (Curtis, 2024). By contrast, we have implemented a two stage strategy in which multiple predictors are used to identify the predictor which potentially is best at recognising pathogenicity for each gene before proceeding to a second stage analysis in a separate dataset for which only that predictor is used to test for association.

The ExWAS did not incorporate APOE status as a covariate in the primary analyses. For some secondary analyses, participants were dichotomised into whether or not they carried an APOE ε4 allele. However the APOE alleles exhibit a dosage effect and additionally the ε3 allele confers excess risk relative to the ε2 allele (Bertram et al., 2007). Hence, we incorporated both ε3 and ε4 doses as covariates in our primary analyses, along with sex and population principal components.

## Methods

Relevant UK Biobank phenotype fields had been downloaded in August 2020. The exome sequence data was released in two phases. Data had been downloaded for 200,632 subjects who had undergone exome-sequencing and genotyping by the UK Biobank Exome Sequencing Consortium using the GRCh38 assembly with coverage 20X at 95.6% of sites on average (Szustakowski et al., 2021). The UK Biobank Research Analysis Platform (RAP) was used to access the Final Release Population level exome variants in PLINK format for 469,818 exomes which had been produced at the Regeneron Genetics Center based on DNA extracted from stored blood samples and using the protocols described here: https://dnanexus.gitbook.io/uk-biobank-rap/science-corner/whole-exome-sequencing-oqfe-protocol/protocol-for-processing-ukb-whole-exome-sequencing-data-sets (Backman et al., 2021). Data from the first cohort of 200,000 participants was used for exploratory first stage analyses and then data from the remaining 270,000 participants was used in confirmatory analyses. UK Biobank had obtained ethics approval from the North West Multi-centre Research Ethics Committee which covers the UK (approval number: 11/NW/0382) and had obtained informed consent from all participants. The UK Biobank approved an application for use of the data (ID 51119) and ethics approval for the analyses was obtained from the UCL Research Ethics Committee (11527/003).

A dementia score was derived from a combination of self report of dementia in one or both parents along with an individual diagnosis of dementia. If a participant self-reported that they had a diagnosis of dementia, if they had been assigned an ICD9 or ICD10 code for dementia or if they reported that both parents had dementia then they were assigned a score of 2. If they had no individual diagnosis of dementia but reported that one parent had dementia they were assigned a score of 1 and otherwise they were assigned a score of 0.

Attention was restricted to variants with minor allele frequency (MAF) < 0.01. All exonic variants were annotated using Variant Effect Predictor (VEP) (McLaren et al., 2016). Nonsynonymous variants were additionally annotated using the AlphaMissense plug-in of VEP, which produces a raw score and a categorisation of likely pathogenic, likely benign or ambiguous, these three categories being converted to numerical scores of 2, 0 or 1 respectively (Cheng et al., 2023). Additionally, rank prediction scores were obtained for an additional 43 different predictors of pathogenicity as provided for all possible nonsynonymous variants in dbNSFP v4 (Liu et al., 2020).

To obtain population principal components reflecting ancestry, version 2.0 of *plink* (https://www.cog-genomics.org/plink/2.0/) was run with the options *--maf 0*.*1 --pca 20 approx* (Chang et al., 2015; Galinsky et al., 2016). For each participant, the number of *APOE* ε3 and ε4 alleles was determined using the genotypes for rs429358 and rs7412.

In order to carry out weighted burden analysis, for each LOF or nonsynonymous variant a score was defined. For LOF variants this score used a parabolic function of allele frequency as previously described, with extremely rare variants (MAF ~ 0) being assigned a score of 10 and less rare variants (MAF=0.01) being assigned a score of 1 (Curtis, 2012, 2022). For nonsynonymous variants, this frequency score was assigned in the same way but was then multiplied by the score for a pathogenicity predictor in order to produce an overall score for that variant. For each gene, each participant would receive a LOF score and a nonsynonymous score, consisting of the sums of the scores for these classes of variant carried by that participant. To test for association, multiple linear regression was performed using sex, 20 principal components and APOE ε3 and ε4 doses as covariates. For the null model, dementia score was predicted solely from these covariates. For the alternative model, dementia score was predicted from these covariates along with the LOF and nonsynonymous scores. The likelihoods of these models were compared and a p value obtained assuming that twice the log likelihood difference followed a chi-squared distribution with two degrees of freedom and the statistical evidence for association was summarised as minus log10(p) (MLP).

Because previous work had shown that LOF variants and nonsynonymous variants could make different relative contributions to disease risk and because different predictors of pathogenicity had differential performance across different genes, a two-stage approach to analysis was followed (Curtis, 2024). In the first stage, the first cohort of 200,000 exome-sequenced participants was used to carry out 45 different regression analyses as described above for each protein-coding gene using each of the 45 pathogenicity predictors. This yielded 45 MLPs for each gene, the maximum being denoted MaxMLP. In the second stage, the remaining 270,000 participants were used for analyses of only the 100 genes showing the strongest evidence for association in the first phase and each gene was analysed using only the predictor which had produced the maximum MLP for that gene in the first phase. Thus only 100 tests were performed in the second phase, meaning that a gene could be declared statistically significant if it produced a test statistic significant at p < 0.05/100.

For each gene achieving statistical significance, the analysis was repeated using the best predictor in all 470,000 participants. Additionally, a full linear regression analysis was carried out in all participants including the same covariates and using the raw counts for all categories of variants as characterised by VEP, along with counts of variants having a dbNSFP rank score for the best performing predictor of more than a threshold value of 0.35. These additional analyses were also carried out for any genes which did not achieve significance but which had been reported to be implicated by the previous ExWAS.

## Results

Out of 469,739 exome sequenced participants with APOE genotypes available, 58,589 were assigned a dementia score of 1 and 6,420 a score of 2. In the first stage analysis, MLPs were obtained for 19,008 protein coding genes using 45 different predictors and these are listed in Supplementary Table 1. The 100 genes with the highest MaxMLPs went forward into the second stage analysis and Supplementary Table 2 shows the MLPs obtained in the second cohort of 270,000 participants using the same predictor as had yielded the maximum MLP in the first 200,000 participants. Of these, three genes reached the threshold for statistical significance, *TREM2, SORL1* and *ABCA7*, and these results are shown in Table 1. As can be seen, all three of these genes produced MLPs over 5 in the second stage analysis, exceeding the critical threshold of −log10(0.05/100) = 3.3. In the full sample of 470,000, these three genes all also produced MLPs which would exceed a conservative threshold for multiple-testing of −log10(0.05/(19008*45)) = 7.23.

**Table 1:** For each gene achieving statistical significance in the second stage analysis, the table shows the maximum −log10(p) (MLP) produced by any of 45 pathogenicity predictors in the first stage analysis of 200,000 participants along with the predictor producing this MLP, results obtained in the second stage using 270,000 participants and in the total sample of 470,000. All results are obtained from multiple linear regression analyses of dementia score with sex, 20 principal components and APOE genotypes as covariates and MAF weighted LOF scores and pathogenicity scores as predictors.

| Gene | MaxMLP200K | Predictor | MLP270K | MLP470K |
| --- | --- | --- | --- | --- |
| *TREM2* | 9.83 | CADD_raw_rankscore_hg19 | 5.04 | 13.14 |
| *SORL1* | 8.94 | Alpha_Missense_score | 5.66 | 14.49 |
| *ABCA7* | 6.62 | LIST_S2_rankscore | 5.26 | 8.75 |

Table 2 shows the results for these genes of analysing the raw counts of variants in all categories assigned by VEP along with variants scoring higher than 0.35 with the relevant predictor. Multiple linear regression analysis to quantify the effect on dementia score was carried out using all variant counts along with sex, 20 principal components and APOE genotypes as covariates. The effect on dementia score does not translate directly to an effect on risk of dementia but for comparison we can note that in these analyses the beta value for APOE ε4 versus ε2 was 0.088 and for ε3 versus ε2 was 0.014 and the corresponding ORs for the effects of these alleles on risk of Alzheimer’s disease (versus ε2) have been estimated to be approximately 5 and 1.7 respectively (Bertram et al., 2007). The beta values for LOF variants in *TREM2, SORL1* and *ABCA7* are 0.191, 0.112 and 0.034. For all these genes the beta values for nonsynonymous variants scoring highly on the relevant predictors are considerably lower, consisting of 0.029, 0.012 and 0.008 respectively, although the value of beta/SE remains high because these nonsynonymous variants are observed more frequently than the LOF variants. Some other variant categories do also produce large values for beta/SE but these are associated with only small effect sizes, with the exception that for *SORL1* 5 prime UTR variants and indels also appear to be associated with a higher dementia score.

**Table 2:** Table showing results of multiple linear regression analysis of dementia score using raw counts of variants in different categories as well as those variants having a score of > 0.35 for the relevant predictor. Analyses were carried out in the full sample of 470,000 participants. Results are shown for all categories for which beta/SE was at least 2. All analyses included sex, principal components and APOE genotypes as covariates. For comparison, the beta value for APOE ε4 versus ε2 was 0.088 and for ε3 versus ε2 was 0.014.

| Gene / Variant category | Number of variants | Variant allele count | Beta (SE) | Beta/SE |
| --- | --- | --- | --- | --- |
| *TREM2* |  |  |  |  |
| LOF | 18 | 64 | 0.191 (0.049) | 3.90 |
| Protein altering | 161 | 21735 | 0.008 (0.003) | 2.31 |
| Synonymous | 69 | 1775 | 0.019 (0.009) | 2.08 |
| CADD_raw_rankscore_hg19 | 73 | 6728 | 0.029 (0.007) | 4.27 |
| *SORL1* |  |  |  |  |
| Five prime UTR | 8 | 65 | 0.105 (0.049) | 2.15 |
| InDel etc | 5 | 17 | 0.324 (0.095) | 3.41 |
| LOF | 101 | 226 | 0.112 (0.026) | 4.36 |
| Protein altering | 1,377 | 117,358 | 0.004 (0.001) | 3.03 |
| Alpha_Missense_score | 461 | 16,621 | 0.012 (0.003) | 3.57 |
| *ABCA7* |  |  |  |  |
| Intronic etc | 2,758 | 3,694,232 | 0.001 (0.000) | 5.02 |
| LOF | 257 | 7,658 | 0.034 (0.004) | 7.59 |
| Protein altering | 1,851 | 1,077,779 | -0.002 (0.001) | -4.33 |
| Splice region | 288 | 461,375 | 0.004 (0.001) | 5.13 |
| Synonymous | 868 | 2,029,732 | -0.002 (0.000) | -5.47 |
| LIST_S2_rankscore | 244 | 66917 | 0.008 (0.002) | 3.68 |

Nine of the genes implicated in the previous ExWAS were not identified by the present analyses and for these genes the MLPs obtained in the 200,000, 270,000 and full dataset are shown in Table 3. It can be seen that for two genes, *GRN* and *PSEN1*, there is modest evidence for association which does not achieve conventional standards of statistical significance after correction for multiple testing. However for the other seven genes the methods used here do not detect any appreciable evidence in favour of association. The results of analyses of raw counts of individual variant categories for these nine genes are shown in Table 4, for LOF variants and all other variant categories for which the magnitude of beta/SE was at least 2. From this it can be seen that LOF variants in *GRN* appear to be associated with quite a strong effect on dementia score but that there is no appreciable signal from other variant categories. The situation is reversed for *PSEN1*, in that LOF variants appear to have negligible effect (with beta = −0.005) while variants with high Alpha Missense scores do seem to have a moderate effect (beta = 0.022), intermediate between the effects of APOE ε3 and ε4 alleles. For other genes there is no evidence for an appreciable effect of any individual variant category because either the estimate of beta is low or because beta/SE is low. Full results for these variant count analyses in all nine genes are shown in Supplementary Table 3.

**Table 3:**
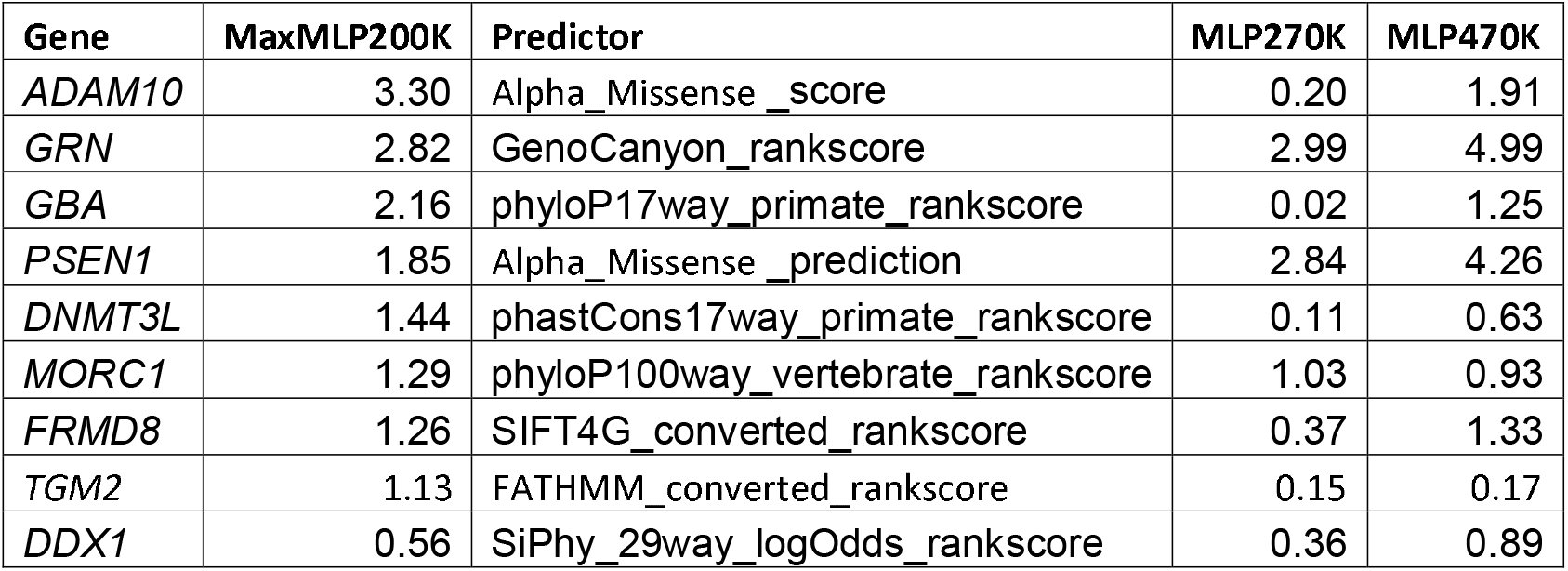
As for Table 1, showing MLPS for the genes which were not significant in the current study but which were implicated by the previous ExWAS.

**Table 4:** As for Table 2, results of multiple linear regression analysis of dementia score using raw counts of variants in different categories for genes implicated by the previous ExWAS but which were not significant in the current study. Results are shown for LOF variants and for all other categories for which the magnitude of beta/SE was at least 2.

| Gene / Variant category | Number of variants | Variant allele count | Beta (SE) | Beta/SE |
| --- | --- | --- | --- | --- |
| *GRN* |  |  |  |  |
| LOF | 52 | 201 | 0.227 (0.028) | 8.18 |
| Synonymous | 216 | 33,016 | 0.006 (0.002) | 2.54 |
| Three prime UTR | 34 | 185,995 | 0.004 (0.001) | 3.48 |
| *PSEN1* |  |  |  |  |
| LOF | 20 | 449 | -0.005 (0.021) | -0.23 |
| Alpha_Missense_prediction | 53 | 135 | 0.022 (0.004) | 6.14 |
| *GBA* |  |  |  |  |
| LOF | 38 | 302 | 0.028 (0.023) | 1.22 |
| Protein altering | 329 | 25,736 | 0.009 (0.003) | 3.37 |
| Three prime UTR | 9 | 3,373 | 0.018 (0.006) | 2.97 |
| *ADAM10* |  |  |  |  |
| Intronic etc | 1107 | 1,023,437 | -0.001 (0.000) | -3.41 |
| LOF | 24 | 44 | 0.057 (0.057) | 1.01 |
| Alpha_Missense_score | 140 | 556 | 0.047 (0.018) | 2.58 |
| *FRMD8* |  |  |  |  |
| LOF | 43 | 182 | -0.017 (0.026) | -0.67 |
| *DDX1* |  |  |  |  |
| Intronic etc | 1332 | 1,929,931 | 0.001 (0.000) | 2.5 |
| LOF | 25 | 54 | 0.019 (0.054) | 0.36 |
| Synonymous | 165 | 387,635 | 0.003 (0.001) | 2.63 |
| *DNMT3L* |  |  |  |  |
| LOF | 24 | 333 | -0.001 (0.022) | -0.04 |
| *TGM2* |  |  |  |  |
| LOF | 50 | 390 | -0.010 (0.020) | -0.48 |
| *MORC1* |  |  |  |  |
| LOF | 51 | 477 | 0.001 (0.016) | 0.07 |

## Discussion

The study provides evidence for association between the familial risk for dementia and rare coding variants in *TREM2, SORL1* and *ABCA7*. Although the target phenotype consisted of undifferentiated dementia, in fact these genes have all been implicated in risk of AD and this study did not identify any genes which might be expected to impact risk of VD (Holstege et al., 2022). The previous ExWAS failed to find significant association with *TREM2*, but did pick up two other known dementia genes, *GRN* and *PSEN1*, both of which have been reported to be associated with risk of AD and of frontotemporal dementia (Curtis et al., 2019; Mendez & McMurtray, 2006; Vardarajan et al., 2022; Zhang et al., 2024). A possible contribution to the current study’s failure to produce strong evidence for association with these genes is that the methodology used assumes that both LOF and nonsynonymous variants contribute to risk and hence implements a two degree of freedom test. However, for both these genes, only one category of variant in fact has an appreciable effect on risk and in this situation using a test with two degrees of freedom is expected to have reduced power.

Two additional genes identified by the ExWAS had previously been reported to be associated with dementia, *GBA* and *ADAM10*, but produce little evidence for association with the methods used in the current study and hence should probably be viewed as false negative results. However, the other five genes implicated by the ExWAS were described as novel and hence the question arises as to whether they are false negative results for this study or false positive results for the ExWAS. Of these one gene, *TGM2*, was not significant in the main analysis at FDR-Q < 0.05 but was implicated only in subgroup analyses of APOE ε4 carriers at p values which would not survive Bonferroni correction for multiple testing (Zhang et al., 2024). Although the authors claim at one point that association with dementia was significant at p < 0.05 for all these five novel genes in the FinnGen study, elsewhere they state they used only single variant results from FinnGen so apparently the claim is that for each gene they could find at least one variant which was significant at p < 0.05 (Kurki et al., 2023; Zhang et al., 2024). Overall, it seems reasonable to conclude that whether rare coding variants in these five genes actually increase dementia risk remains somewhat open to question.

Our results provide useful information about the nature of the effect variants in the implicated genes have on dementia risk. LOF variants in TREM2 and SORL1 have effect sizes greater than the APOE ε4 allele but are extremely rare and are cumulatively seen in only around 1/2000 people. By contrast, LOF variants in ABCA7 have an effect intermediate between APOE ε3 and ε4 alleles and are much less rare, being carried by around 1/70 people. Nonsynonymous variants in these genes with a high score for a pathogenicity predictor are likewise not very rare and have effect sizes comparable to the effect of ε3 (compared to ε2). Importantly, the relevant pathogenicity predictor is different for each gene, being CADD for *TREM2*, AlphaMissense for *SORL1* and LIST-S2 for *ABCA7* (Cheng et al., 2023; Malhis et al., 2020; Rentzsch et al., 2019). If novel therapies become available which can prevent the development of Alzheimer’s disease in those at risk then it will be helpful to accurately evaluate genetic risk and to use the most appropriate pathogenicity predictor for each gene in order to do so.

Studies such as this may throw further light on the mechanisms by which genetic variation can influence disease risk. For example, there has recently been controversy about the role of TREM2 in the pathogenesis of AD and whether higher levels of soluble TREM2 might be associated with increased or decreased disease progression (Pomara & Imbimbo, 2024; Wagemann et al., 2024). Our results make very clear that LOF variants in *TREM2*, expected to lead in to reduced activity, are associated with higher risk of dementia. TREM2 modifies microglial function in the context of neurodegeneration, possibly via the mammalian target of rapamycin (mTOR) pathway, and may affect microglial responses to Aβ (Ulland et al., 2017; Wang et al., 2020). SORL1 is a receptor involved in the endocytic pathway which contributes to the generation of Aβ through the re-entry and recycling of APP from the cell surface. Overexpression of *SORL1* results in decreased Aβ production while suppression results in overproduction of Aβ and it has been proposed that SORL1 regulates sorting of APP into the retromer-recycling pathway (Rogaeva et al., 2007). ABCA7 is involved in membrane transport and has also been implicated in the APP processing pathway, while loss of ABCA7 has been shown to aggravate amyloid plaque burden in a mouse model of AD (Kim et al., 2013). While the association of LOF variants in *TREM2, SORL1* and *ABCA7* with increased dementia risk confirms that reduced activity of these genes in some way promotes disease activity, it is possible that further insights into the exact mechanisms involved could be achieved by more detailed study of the positions and nature of the nonsynonymous variants most associated with risk. This will be the subject of future work.

This study further characterises the contribution which rare coding variants can make to risk of dementia in the general population. It produces results which are discrepant from a similar analysis of the same dataset, in that it successfully identifies *TREM7*, which was not picked up by the previous study, but fails to identify other genes known to be involved in dementia susceptibility. These discrepancies highlight that we do not yet have consistently optimal schemes for analysing rare coding variants and exploring improved methodologies remains worthwhile.

## Data availability

The raw data is available on application to UK Biobank at https://ams.ukbiobank.ac.uk/ams/. Derived variables will be returned to UK Biobank, from which they will become available in due course. Software and scripts used to perform the analyses is available at https://github.com/davenomiddlenamecurtis.

## Acknowledgments

This research has been conducted using the UK Biobank Resource under Application Number 51119. The authors wish to acknowledge the staff supporting the High Performance Computing Cluster, Computer Science Department, University College London. The authors wish to thank the participants who volunteered for the UK Biobank project. This work uses data provided by patients and collected by NHS England as part of their care and support. This research also used data assets made available by National Safe Haven as part of the Data and Connectivity National Core Study, led by Health Data Research UK in partnership with the Office for National Statistics and funded by UK Research and Innovation (grants MC_PC_20029 and MC_PC_20058). For the work to carry out the analyses reported here, the authors did not receive any specific grant from funding agencies in the public, commercial, or not-for-profit sectors.

